# Model-based Bayesian inference of the ventilation distribution in patients with Cystic Fibrosis from multiple breath washout, with comparison to ventilation MRI

**DOI:** 10.1101/2021.10.01.21264402

**Authors:** Carl A. Whitfield, Alexander Horsley, Oliver E. Jensen, Felix C. Horn, Guilhem J. Collier, Laurie J. Smith, Jim M. Wild

## Abstract

**Background:** Indices of ventilation heterogeneity (VH) from multiple breath washout (MBW) have been shown to correlate well with VH indices derived from hyperpolarised gas ventilation MRI. Here we report the prediction of ventilation distributions from MBW data using a mathematical model, and the comparison of these predictions with imaging data.

**Methods:** We developed computer simulations of the ventilation distribution in the lungs to model MBW measurement with 3 parameters: *σ*_*V*,_ determining the extent of VH; *V*_0_, the lung volume; and *V*_*D*_, the dead-space volume. These were inferred for each individual from supine MBW data recorded from 25 patients with cystic fibrosis (CF) using approximate Bayesian computation. The fitted models were used to predict the distribution of gas imaged by ^3^He ventilation MRI measurements collected from the same visit.

**Results:** The MRI indices measured (*I*_1/3_, the fraction of pixels below one-third of the mean intensity and *I*_*CV*_, the coefficient of variation of pixel intensity) correlated strongly with those predicted by the MBW model fits (*r* = 0.93, 0.88 respectively). There was also good agreement between predicted and measured MRI indices (mean bias ± limits of agreement: *I*_1/3_ : − 0.003 ± 0.118 and *I*_*CV*_: − 0.004 ± 0.298). Fitted model parameters were robust to truncation of MBW data.

**Conclusion:** We have shown that the ventilation distribution in the lung can be inferred from an MBW signal, and verified this using ventilation MRI. The Bayesian method employed extracts this information with fewer breath cycles than required for LCI, reducing acquisition time required, and gives uncertainty bounds, which are important for clinical decision making.

## 1. Introduction

Ventilation heterogeneity (VH) refers to the unevenness of inspired air distribution in different lung regions during breathing. It is an early and prominent feature of lung diseases such as cystic fibrosis (CF), bronchiectasis, asthma and COPD^1–4^. Clinical assessment of VH in the lung is performed by the multiple breath washout test (MBW), from which the most commonly used primary outcome is the lung clearance index (LCI)^5^. LCI is now well established, particularly in CF where it is a sensitive, robust measure of early disease, and responsive to clinical status^6^. However, LCI utilises a relatively small proportion of the gas washout data collected: the alveolar gas concentrations, and specifically those preceding the first and last washout breaths. Alternative VH indices from MBW, such as moment ratios^7^ and phase-III slopes^8^, have been developed but are less commonly used as they are more sensitive than LCI to other factors besides ventilation heterogeneity^5,9^, including variations in tidal volume and gas diffusivity. One current limitation to widespread clinical use of LCI as a pulmonary function test is the time required for data collection^5^. Although LCI appears robust to earlier test thresholds to some extent, this may be at the cost of reduced sensitivity and the practice is not widespread^10–14^.

Hyperpolarised gas ventilation MRI allows for visualisation of the ventilation distribution in the lung^15^. Patients inhale a bolus of hyperpolarised tracer gas (^129^Xe or ^3^He) which is imaged during a short breath hold (as in this study), dynamically during the respiratory cycle^16^ or over several cycles ^17^. This enables the identification of small ventilation defects that are not detected by spirometry (FEV_1_) or LCI, and the method is therefore highly sensitive to early lung disease progression^1,18,19^. Ventilation MRI also provides a 3-dimensional representation of the distribution of lung disease, allowing regional changes to be identified and tracked. Furthermore, MRI can identify lung regions where tidal flow is entirely obstructed, which would not normally contribute to the MBW signal. This powerful technique is therefore potentially more informative about the nature and severity of lung disease. We have previously reported a good correlation between MRI markers of airway obstruction and LCI in patients with CF^18,20^. The availability of these paired data presents a unique opportunity to link clinically usable measures of gas washout with detailed lung imaging in order to better inform understanding of MBW and to develop more sophisticated washout metrics.

To improve the clinical viability of MBW, we have developed a Bayesian method for inferring the distribution of ventilation directly from MBW data. Underlying the method is an efficient three-parameter model of gas ventilation and transport in the lung similar to previous models in this area. The benefit of the method presented here is that uncertainty in the predictions is readily quantified by the Bayesian methodology, and where there are multiple viable solutions all are given weight relative to their probability of explaining the observed data. The model predictions have been compared to independent measurements of the VH from ventilation MRI, directly testing the validity of the inferred parameters and the predicted ventilation distribution.

The aim of this study was to develop computational software to predict the ventilation distribution in individual subjects using raw MBW data and to use this to produce robust indices of VH that directly correspond to those measured directly with ventilation MRI. A secondary aim was to test whether the measures derived using this new method were robust at shorter test times.

## 2. Methods

Table 1 introduces the mathematical symbols that are used in this section prior to their first use in the text.

**Table 1:** A list of all mathematical symbols and notation used in the text, and what they represent.

| Symbol(s) | Description of use |
| --- | --- |
| $i$ | Index of lung units and dead space compartments in the compartmental lung model. |
| $j$ | Index of volume elements in the compartmental lung model. |
| $k$ | Index of breath number in processed MBW data. |
| $n$ | Time-step index in processed MBW data. |
| $s$ | Voxel index in MRI data (simulated or observed). |
| $I_{1/3}$ | Proportion of the masked $\text{He}^3$ MRI image with voxel brightness less than 1/3 of the mean. |
| $I_{\text{CV}}$ | Coefficient of variation in brightness of masked $\text{He}^3$ MRI image. |
| $\text{LCI}_z$ | Lung clearance index measured by MBW (mean from 3 test repeats), $z = 2.5, 5, 10, 20, 40$ is the termination threshold used in % of equilibrium $\text{SF}_6$ concentration. |
| $V_{\text{FRC}}$ | Functional residual capacity (L) measured by MBW (mean from 3 test repeats). |
| $V_{\text{FDS}}$ | Fowler dead-space (L) measured from $\text{CO}_2$ curve during MBW (mean from 3 test repeats – each test is the median value of all washout breaths). |
| $V_{\text{FRC}}^{(\text{pleth})}$ | Functional residual capacity (L) measured by plethysmography |
| $\tilde{V}_0, \tilde{V}_D$ | Initial guess for model parameters $V_0$ and $V_D$, used to set model priors. |
| $V_k^{(\text{inh})}, V_k^{(\text{exh})}$ | Volume of inhalation and exhalation respectively for breath $k$ of an MBW test. |
| $N_{II}, N_{III}$ | Number of data-points used to represent phase-II and phase-III parts of each breath ($N_{II} = N_{III} = 4$ used throughout). |
| $\{V_0, V_D, \sigma_v\}$ | Compartmental lung model parameters setting the FRC, dead-space volume and VH respectively. |
| $\{v_{ij}, c_{ij}^{(p)}, c_{ij}^{(d)}, g_{ij}\}$ | Properties of the volume element $j$ in dead-space compartment $i$: gas volume, inert gas concentrations (distal end and proximal end), and inert gas volumes respectively. |
| $\{C_i, V_i, G_i\}$ | Properties of the lung unit $i$: inert gas concentration, gas volume and inert gas volume respectively. |
| $N_c$ | Number of lung units in the model ($N_c = 50$ used throughout) |
| $v_{\text{max}}$ | Re-discretisation volume used for mixing-step ($v_{\text{max}}$ = used throughout) |

### 2.1 Study design and recruitment

As part of a longitudinal observational study, children and adults with CF were recruited from three UK specialist centres^19,20^. At recruitment, patients had to be over the age of five years, be clinically stable for four weeks prior to their visit and achieve an FEV_1_ >30% predicted within the previous six months. This study was approved by the Yorkshire and Humber - Leeds West Research Ethics Committee (REC reference: 16/YH/0339). Parents/guardians of children and all adult patients provided written informed consent. For this analysis we have used data from a single visit when patients completed both MRI and MBW, including supine MBW (since this corresponds to the same position as the MRI procedure, and so minimises the effect of changes due to body position or gravity^21,22^).

### 2.2 Ventilation MRI

Ventilation MRI was performed on a 1.5T GE HDx scanner (GE, Milwaukee, WI, USA) using hyperpolarised helium-3 (^3^He) using a transmit-receive vest coil (CMRS, Milwaukee, WI, USA) and a three-dimensional (3D) steady state free precession ventilation imaging sequence as described previously^17^. Images used in this study were acquired during a breath-hold at end-inspiratory tidal volume following the inhalation of a predetermined fixed volume of test gas from their resting functional residual capacity (FRC). The volume of gas was titrated based on the subject’s height and consisted of scaled doses of ^3^He balanced with nitrogen^20^.

Contiguous ventilation MR images of the coronal plane were acquired with slice thickness of 5mm and pixel size ranging from 2.73 × 2.73mm to 3.28 × 3.28 mm (depending on patient lung size). For each slice, a mask was manually derived from ^1^H images (acquired during the same breath hold as the ^3^He MRI) to determine which pixels corresponded to positions inside the lung cavity (excluding visible airways). The masked images were eroded by one pixel to avoid edge effects. The intensity (brightness) of each pixel is taken as a relative measure of the gas concentration at that point. This is used as a proxy for the local ventilation rate, and is used to characterise VH in two ways (see figure 1):

- Measuring the ‘poorly ventilated’ fraction of the lung by *I*_1/3_. That is, the fraction of pixels with intensity less than one-third of the mean (as used previously to define ventilation defects^23^).
- Calculating the total coefficient of variation of the normalised pixel intensity, *I*_*CV*_. Note these are measures of global ventilation heterogeneity which can be predicted from MBW data, other common MRI indices such as *CV*_mean_^24^ or Δ*R*^25^ quantify spatial heterogeneity and are not considered here.

**Figure 1:**
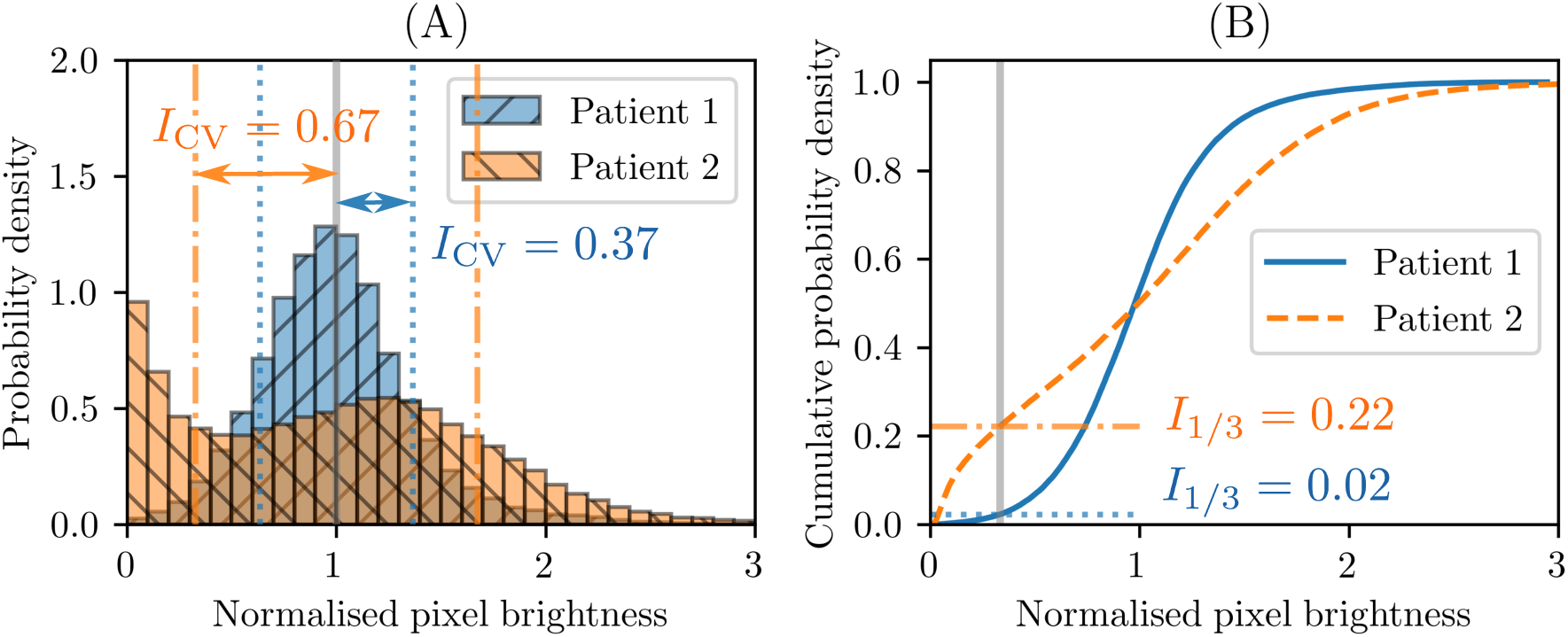
Examples of MR indices I_1/3_ and I_CV_ for two patients. Patient 1 (blue) has low VH, whereas patient 2 (orange) has high VH. (a) Histogram of pixel intensity (normalised to have unit mean, as indicated by the vertical thick grey line). The blue dotted and orange dot-dashed lines show 1 standard deviation from the mean for patient 1 and 2 respectively. The standard deviations of this normalised distribution are the I_1/3_ values (as given in the figure). (b) The cumulative distributions of pixel intensity for the same two patients. Where these curves cross 1/3 of the mean pixel intensity (indicated by the thick grey line) corresponds to the I_1/3_ value on the vertical axis (the blue dotted line for patient 1, and orange dot-dashed line for patient 2), as this is the proportion of the distribution below this value.

### 2.3 Pulmonary function tests

MBW was performed using a modified open-circuit Innocor device (Innovision, Glamsbjerg, Denmark)^26^ using 0.2% sulphur hexafluoride (SF_6_) tracer gas in air. MBW tests were collected in triplicate in the supine position. Spirometry and body plethysmography were performed to international standards^27,28^ using a PFT Pro (Vyaire, Basingstoke, UK). All tests were performed on the same day. Either MBW or MRI was performed first, followed by the other. Spirometry was always performed last to minimise the influence of VH redistribution due to a forced manoeuvre.

MBW data were analysed using a software package for Igor Pro v6 (Wavemetrics Inc., Lake Oswego, OR, USA) as previously described^12^. We extracted from this analysis the following parameters for each patient: LCI_2.5_, lung volume at FRC (*V*_*FRC*_), and Fowler dead-space from CO_2_ curves (*V*_*FDS*_)^29^. We also re-computed LCI for different termination thresholds (5%, 10%, 20%, 40% of initial concentration).

### 2.4 Compartmental lung model

Figure 2 provides schematic overview of the model-fitting process, which this section describes in detail.

**Figure 2:**
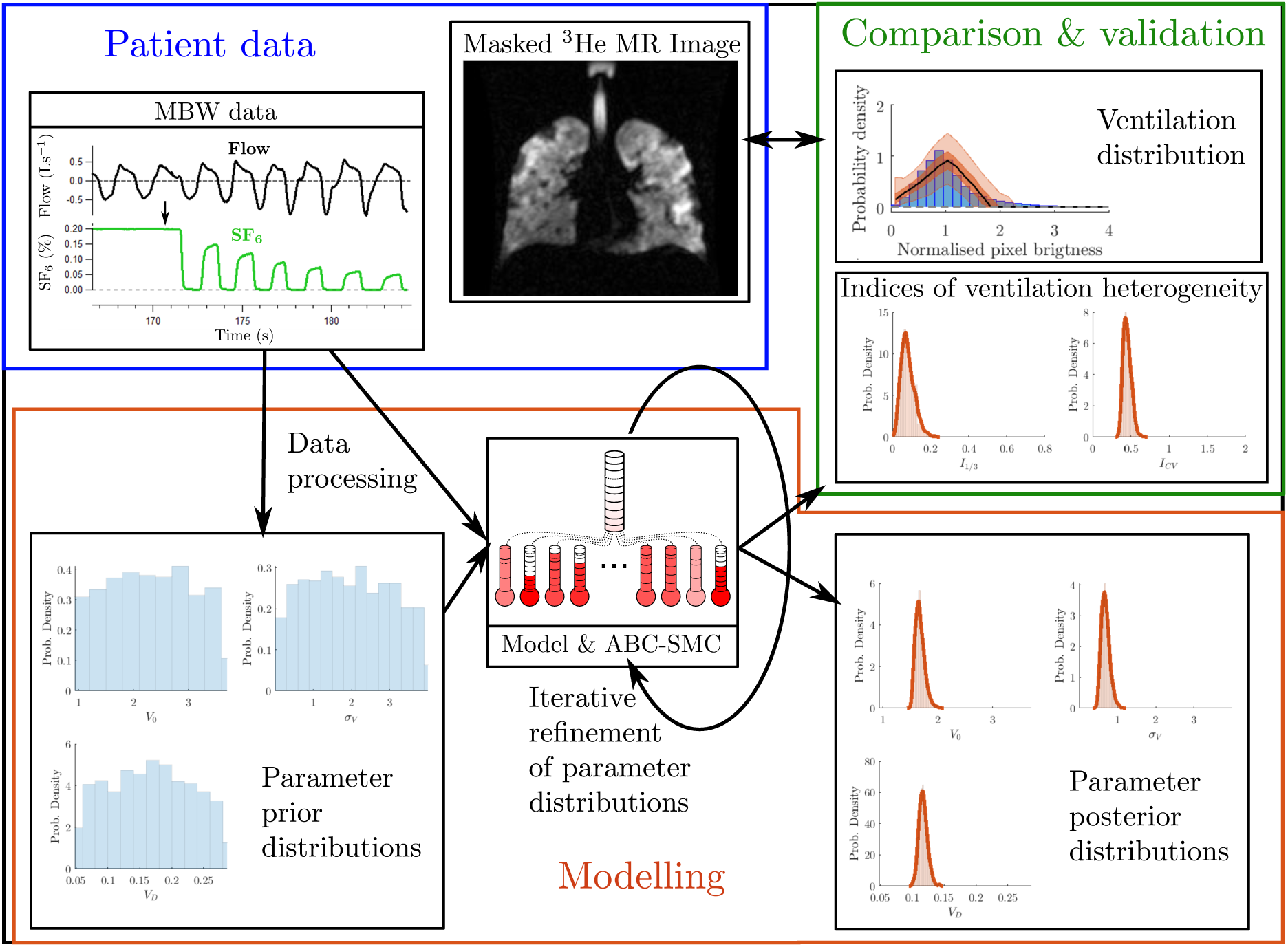
Sketch of the relations between the multiple breath washout (MBW) data, Bayesian (ABC-SMC) model, and imaging data. The raw MBW data (top-left) consist of flow rate (black line), SF6 trace (green line), and CO2 trace (not shown), where the black arrow in the plot indicates switch of gas source (from wash-in to washout). This is processed and split into fewer measurement points in order to be fitted by the model. The model is fitted to the processed data by searching over 3 parameters for each individual. Initially, parameter values are drawn from the prior distributions (blue histograms in bottom-left); the outcome of the ABC algorithm is the posterior distribution of parameters shown in orange in the bottom-right. In both cases the solid lines show a kernel density estimation (KDE) of the distribution, and the most likely parameters (maximum a posteriori) were taken as the peaks of the KDE curves for the posterior distributions. The accepted simulations are also used to predict the probability distribution of ventilation MRI indices I_1/3_ and I_CV_ (middle-right), as well as the ventilation distribution (expressed as a probability density function in the top-right, black line is the median, the dark orange region is the interquartile range, and pale orange the central 95%). The outcomes are then compared to the 3He MRI data (top-centre) where the blue histogram (top-right) shows the normalised probability distribution function of masked image intensity, which in this example closely matches the model prediction.

#### 2.4.1 Processing of MBW data for model fitting

Raw MBW data were outputted as plain text files. Processing of these raw data was carried out using a custom-built C++ program in line with recommendations^5^. Corrections for re-breathed SF_6_ were not applied as these are simulated in the model. In summary:

1. Gas traces from each MBW are corrected for the flow-gas delay (as measured during device calibration).
2. The data are separated into inhalations and exhalations, with a filtering step to ensure that flow fluctuations near to zero are not counted as separate breaths.
3. Exhaled volumes are corrected to body temperature, pressure, water vapour saturated (BTPS) by multiplying the measured flow by 1.016. Then, inhaled volumes are all scaled by a constant factor to give unitary respiratory quotient for the whole test.
4. Fowler dead space (FDS) volume was measured using the CO_2_ traces, and functional residual capacity (FRC) approximated as outlined in Robinson et al. 2013^5^. The median FDS over all valid breaths in each test was used as the test average, then 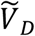 was taken as the mean of this over all the test repeats of an individual. Similarly, 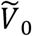 was the mean FRC measured in this step over all test repeats.
5. Finally, exhalations are down-sampled to 10 points per breath: these were the start and end points, 4 evenly spaced points in phase II (defined as exhaled volume between 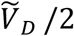 and 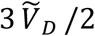), and 4 in phase III (defined as between 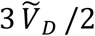 and 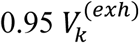), where 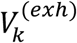 is the total exhalation volume of breath *k*. This down-sampling was achieved by linearly interpolating from the nearest concentration measurements in the dataset. Inhalations were down-sampled to a single step between inhalations, as the inspired gas trace is approximately zero during washout and therefore not used in the fitting process. We label the time-steps with index *n* and the associated volume change as 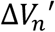.

#### 2.4.2 Compartmental lung model

As in Bates and Peters^30^, the computational lung model comprises *N*_*c*_ = 50 lung units of equal size with total volume *V*_0_ at FRC, with each compartment connected to independent dead-space compartments of equal size with total volume *V*_*D*_. As in Mountain et al.^31^, the relative inflation rate *x* of each compartment is drawn from a lognormal probability distribution with unit mean

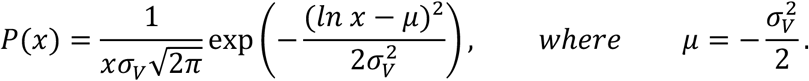

where *σ*_*V*_ is the VH parameter. Once the *x* values have been drawn (*x*_*i*_ *∼ P*(*x*) for *i* = 1,.., *N*_*c*_), they are normalised so that the mean is 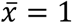. The model simulates advection of the inhaled/exhaled volumes measured from MBW through the dead-space and into/out of the lung units via the transport of discrete volume elements. These volume elements therefore effectively form a Lagrangian grid for the 1D network of dead-space components. Each element *j* is indexed sequentially from distal to proximal end in each dead-space compartment *i* = 0, … *N*_*c*_ (where *i* = 0 is the common-dead space and *i* = 1,.., *N*_*c*_ the private dead-space compartments corresponding to the acinar units with the same index). An element has volume *v*_*ij*_, and concentration values 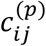 and 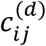 defined at its proximal (mouth) and distal (acinar) ends respectively. These elements are shifted along each time-step by the volume inhaled proximal to them, or exhaled distal to them, as shown in figure 3. A more detailed description of the simulation of each timestep is given in Supplementary Text S1.

**Figure 3:**
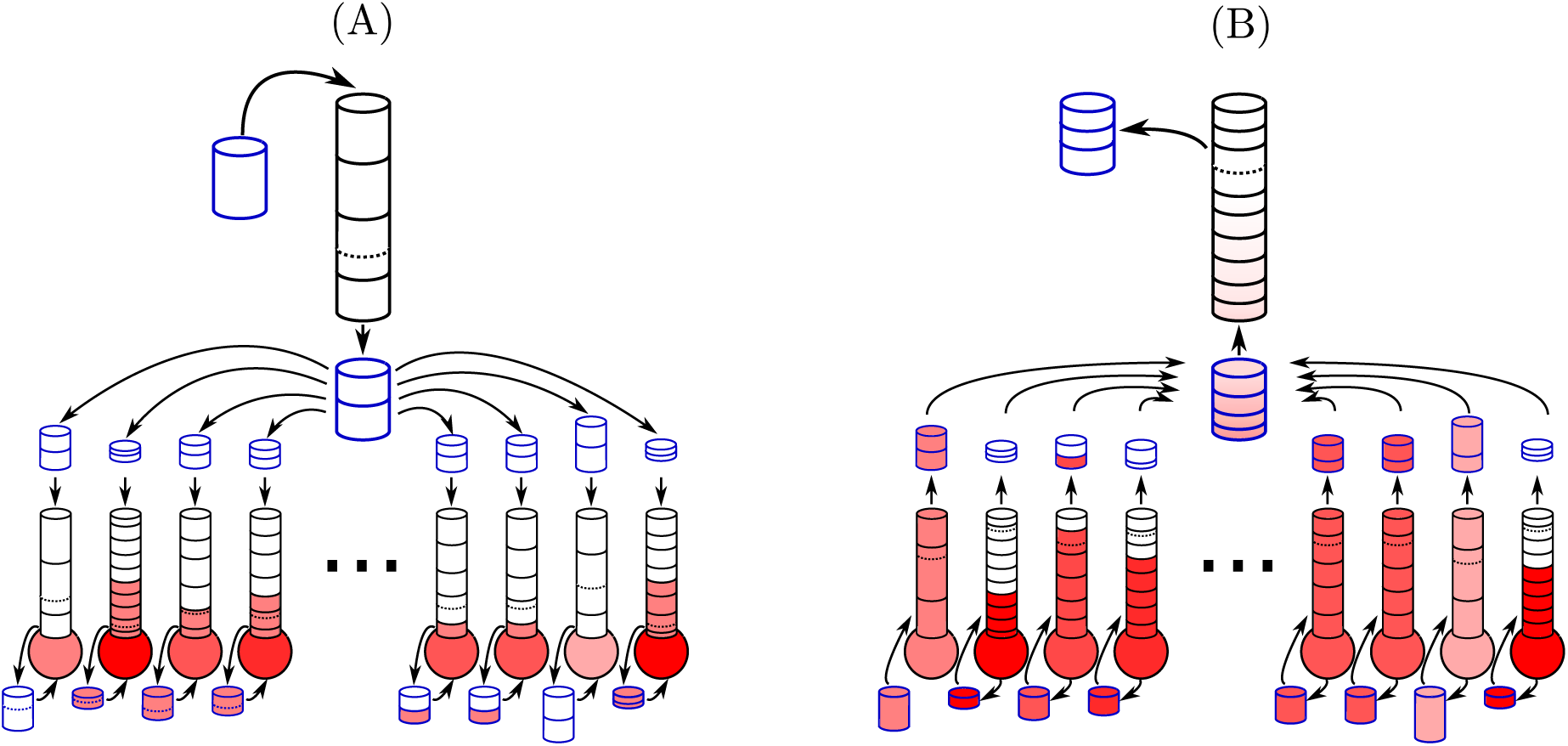
Sketch of a single simulation step where the cylinders represent volume elements inside dead-space compartments, and the spheres represent lung units. The shading in red indicates the concentration of SF_6_ gas. (a) On inhalation a new element (blue outline) is added to the proximal end (top) of the common dead space, and the same amount of volume is removed from the distal end (bottom), as indicated by the dashed black line. This removed volume (blue) is split into volumes for each private dead-space based on their ventilation rate. These are added to the proximal end of each dead-space and the same volume is removed from the distal end (dashed line). This removed gas volume (blue) is then added to the volume of gas in the lung units. (b) On exhalation, the reverse process happens, and there is a re-discretisation of the volumes that are extruded from the private dead-spaces at the mixing point where they are combined.

This model has a number of shared features with previous methods for similar applications, as cited above. It is a stochastic model so simulations with the same distribution parameter (*σ*_*v*_), will have slightly different ventilation distributions, as in Mountain et al. ^31^. The number of compartments in the model (*N*_*c*_) dictates how similar an individual realisation of the ventilation distribution is to the lognormal distribution used to generate it.

#### 2.4.3 Model parameter estimation

Adaptive Bayesian Computation Sequential Monte Carlo (ABC-SMC)^32^ was used for each individual’s data. The flow data from MBW is taken as a model input, and the SF_6_ concentration is the output to be fitted against. Parameters sets are drawn and evaluated to build a (posterior) probability distribution for their actual values through an iterative refinement process, as previously described ^32^ and detailed in Supplementary Text S2.

The parameter prior distributions we assumed uniform and independent as:

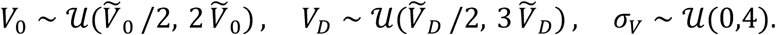

The range bounds are defined using their estimated quantities from MBW processing (see 2.4.1) where 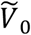 is the estimated FRC (mean of three tests) and 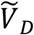 the test median Fowler dead-space (mean of three tests), whereas *σ*_*V*_ is given a broad plausible range.

The posterior parameter sets are used to predict the alveolar ventilation distribution measured by imaging (as detailed in the previous section) and compared directly to those measurements in the same patient. The final number of iterations required in the ABC-SMC algorithm was set adaptively (see Supplementary Text S2 for full details).

### 2.5 Statistical Analysis

Data were analysed in Matlab (v. R2020a)^33^. The MAP values for parameters were estimated using kernel density estimation (KDE) on the posterior parameter distributions (see figure 2 for an example). The KDE used 1000 points with default Matlab settings for the bandwidth, the maximum value was then interpolated by fitting a quadratic function to a 5-point stencil around the maximum of the discretised function and solving for the point where its gradient was equal to 0.

The Pearson correlation *r* was used to quantify correlation between measured and predicted values of the same quantities. Agreement of these values was measured by Bland-Altman analysis. Variability in MBW derived indices (*V*_*FRC*_ and LCI) was quantified by the standard deviation over 3 tests. To compare measures of different properties we have used Spearman’s rank correlation coefficient *ρ*, these are labelled as such in the figures. A p-value of <0.05 was considered statistically significant.

## 3. Results

### 3.1 Parameter identifiability and convergence of the fitting model

We varied the discretisation parameters *N*_*II*_, *N*_*III*_ and *N*_*c*_ to test for convergence of the inferred parameters to those used to generate artificial data from a single model realisation. The results are given in Supplementary Figures S1, S2 and S3. The parameters chosen to fit the data, given in table 1, represent the optimum balance between accuracy and performance.

Supplementary figure S3 shows the parameter recovery from simulated data. We see that, for the full range of *σ*_*V*_ tested (up to 1.5), this heterogeneity parameter is recovered well (within the uncertainty limits). However, we observe that the volume parameters are poorly estimated at very high VH.

### 3.2 Patient population

Paired MRI and LCI data were available for 25 children and adults with CF, with FEV_1_ z-scores ranging from -5.32 to 1.10, and LCI ranging from 6.8 to 16.8. Table 4 shows the patient demographics and lung function of all subjects.

### 3.3 Prediction of physiological parameters and MRI-measured ventilation distribution from MBW data

Figures 4(a) and (c) compare the measured *I*_1/3_ and *I*_*CV*_ values with those predicted from simulations fitted to the MBW data. There is a strong correlation for both measures (*r* = 0.93 and *r* = 0.87 respectively). The Bland-Altman analysis of the predictions vs. measurements in figures 4(b) and (d) shows negligible mean biases of the two measures:−0.003 ± 0.118 and −0.004 ± 0.298 respectively (± 2 S.D.). The uncertainty approximation appears to explain most but not all of the prediction error: for *I*_*CV*_, 18 (72%) measured values fell within the predicted 95% prediction intervals, whilst for *I*_1/3_ it was 20 (80%).

**Figure 4:**
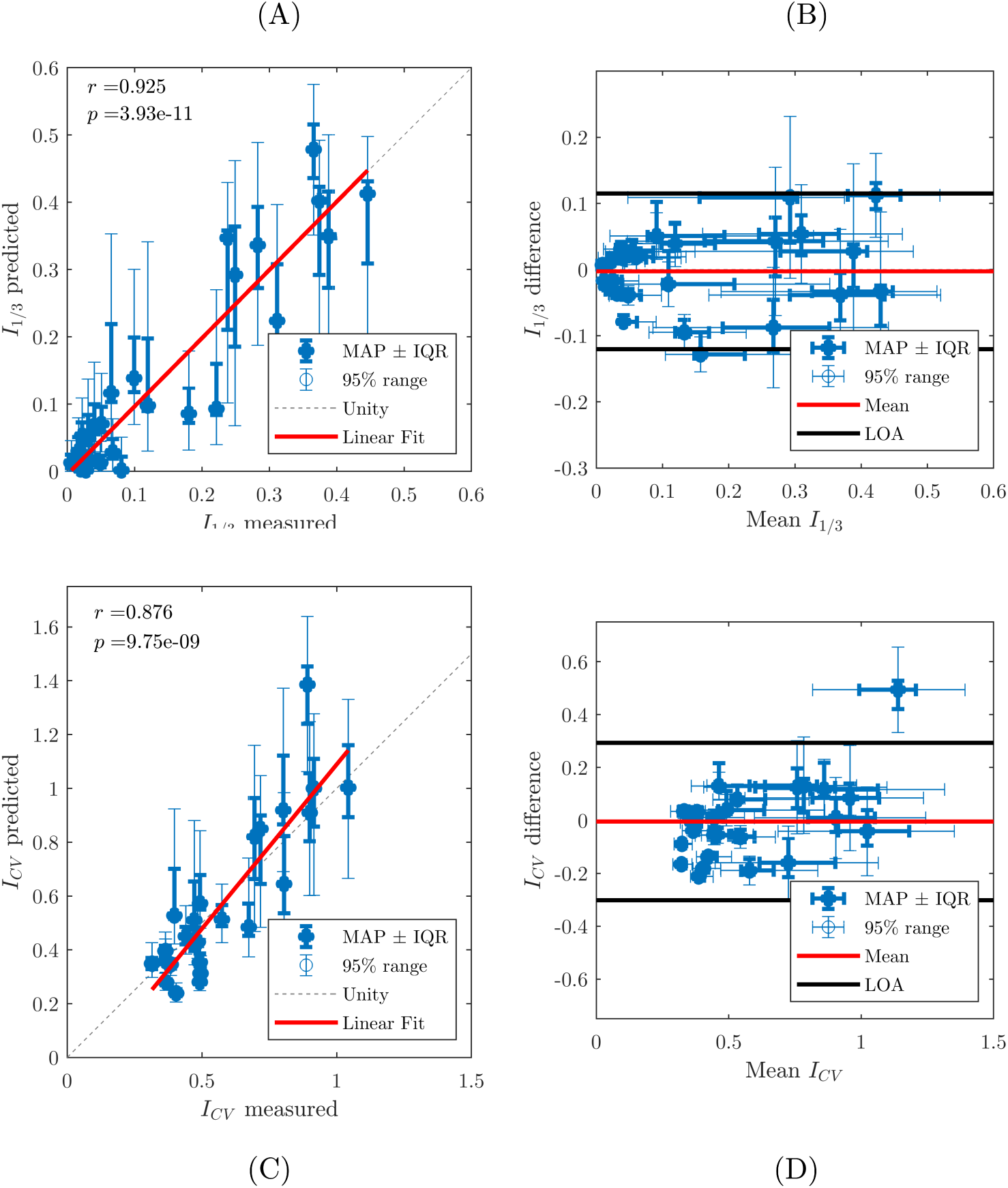
(a) Model predicted I_1/3_ versus MRI measured values. (b) Bland-Altman plot of I_1/3_ (predicted minus measured versus the mean of the two). (c) Model predicted I_CV_ versus MRI measured values. (d) Bland-Altman plot of I_CV_ (predicted minus measured versus the mean of the two). Key: MAP = Maximum a posteriori (most likely measurement value from ABC-SMC algorithm), IQR = Interquartile range (central 50% of sampled posterior distribution from ABC-SMC algorithm), 95% range (central 95% of sampled posterior distribution from ABC-SMC), and LOA = limits of agreement (2 standard deviations either side of the mean).

Supplementary figure S4 compares the posterior model parameters to their MBW counterparts. Figures S4(a) and (b) show that the parameters *V*_0_ and *V*_*D*_ agree well with the MBW measured values *V*_*FRC*_ and *V*_*FDS*_ in the majority of cases (r=0.88 and 0.76 respectively). Figure S4(c) shows that there is strong correlation between the VH parameter *σ*_*V*_ and LCI measured by MBW (r=0.93).

Furthermore, the fitted parameters of the ventilation model show interdependence. Figures S5(a) and (b) shows that increased ventilation heterogeneity *σ*_*V*_ results in an increase of the fitted FRC volume (*V*_0_) and dead-space volume (*V*_*D*_) relative to the value directly computed by MBW. These differences are due to the assumptions underlying both the model and the MBW measures themselves, which break down at high VH, as explained in the discussion. Supplementary figure S5(c) also shows that the uncertainty in predictions increases with VH.

In the majority of cases, the imaged ventilation distributions show good agreement with those predicted by the model (Figure 5). These fits are quantified in Supplementary Table S1, where it is shown that they fit the ventilation distributions better than a single parameter distribution fit directly to the imaging data. Supplementary figure S6 shows two examples of fitted SF_6_ curves compared to MBW data.

**Figure 5:**
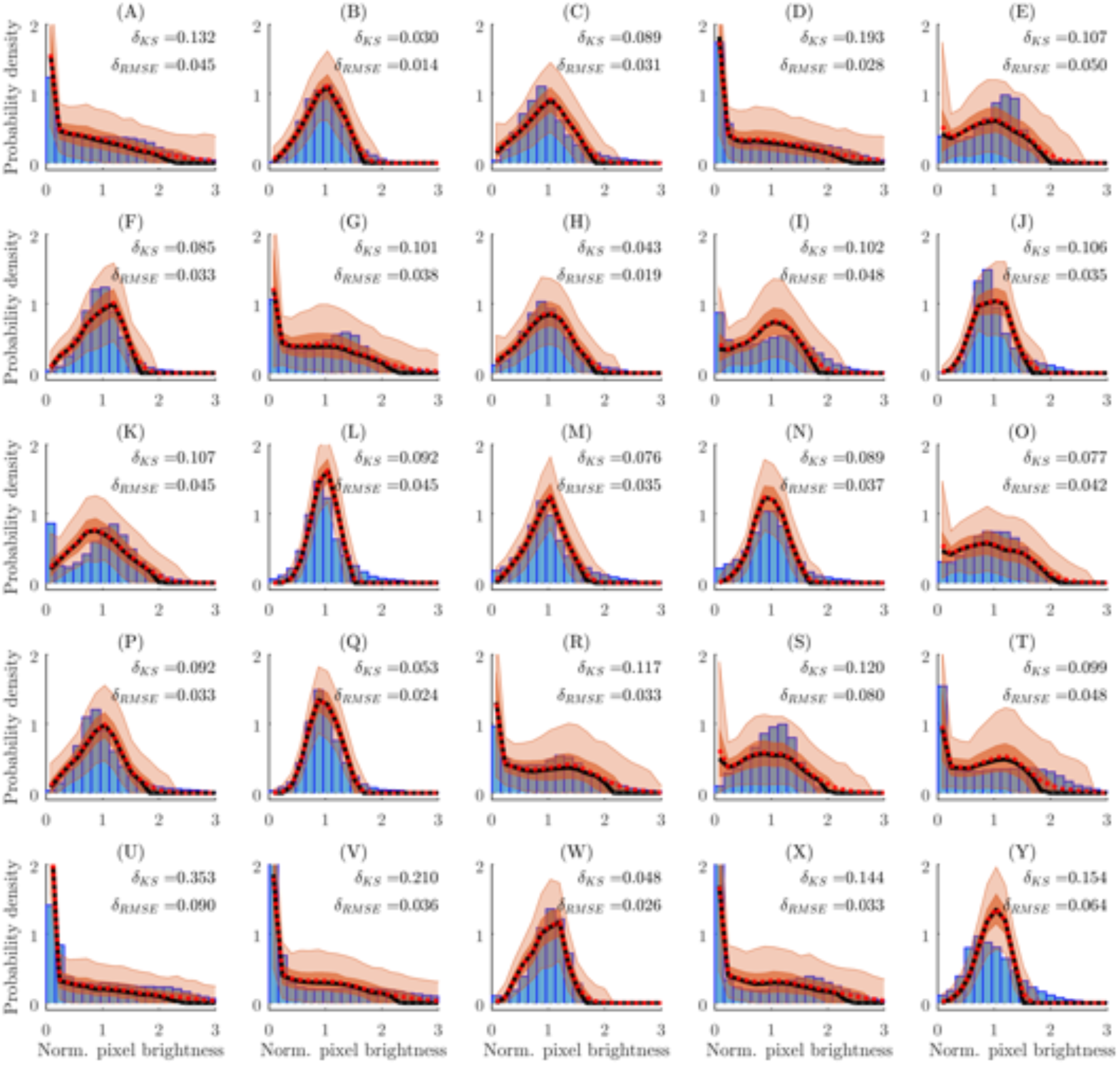
Comparison of MRI measured ventilation distributions (blue histograms, normalised by mean pixel intensity in lung region) against those predicted by modelling. The black line shows the median, the darker orange region the interquartile range, the light orange region the central 95%, and the red dotted line the mean of the MRI ventilation distributions predicted from the final set of accepted simulations from ABC-SMC for each histogram. In the inset of each graph is printed the Kolmogrov-Smirnov (K-S) statistics for both the median and mean predicted distributions (vs. the observed distribution). A bin-width of 0.16 was used to visualise these distributions. Each figure (a)-(y) shows the result for one of the 25 patients in this study.

### 3.4 Impact of shorter washout

To measure the effect of shortening the MBW test time, we computed the LCI and model fits for increasing MBW termination thresholds (retaining the same model priors for *V*_0_ and *V*_*D*_). As the termination threshold is raised, LCI correlates strongly with MRI indices for thresholds of 10% and below (table 5), but this correlation drops off rapidly for larger thresholds.

Figure 6 also shows that LCI values correlate much less strongly with LCI_2.5_ values as the threshold is increased, however the model parameter σ_*V*_ remains practically unchanged up to the 20% threshold, and still correlates strongly with its initial values even at 40% termination threshold (r=0.91 for σ_*V*_ vs. r=0.69 for LCI). As shown in table 3, the correlation of σ_*V*_ with MRI indices of VH (*I*_1/3_ and *I*_*CV*_) is also better maintained at the 20% threshold compared to LCI. Agreement of the model predicted MRI parameters is just as strong at 20% termination threshold as it is at 2.5%. Two examples of model fits for increasing threshold are given in Supplementary figure S7.

**Figure 6:**
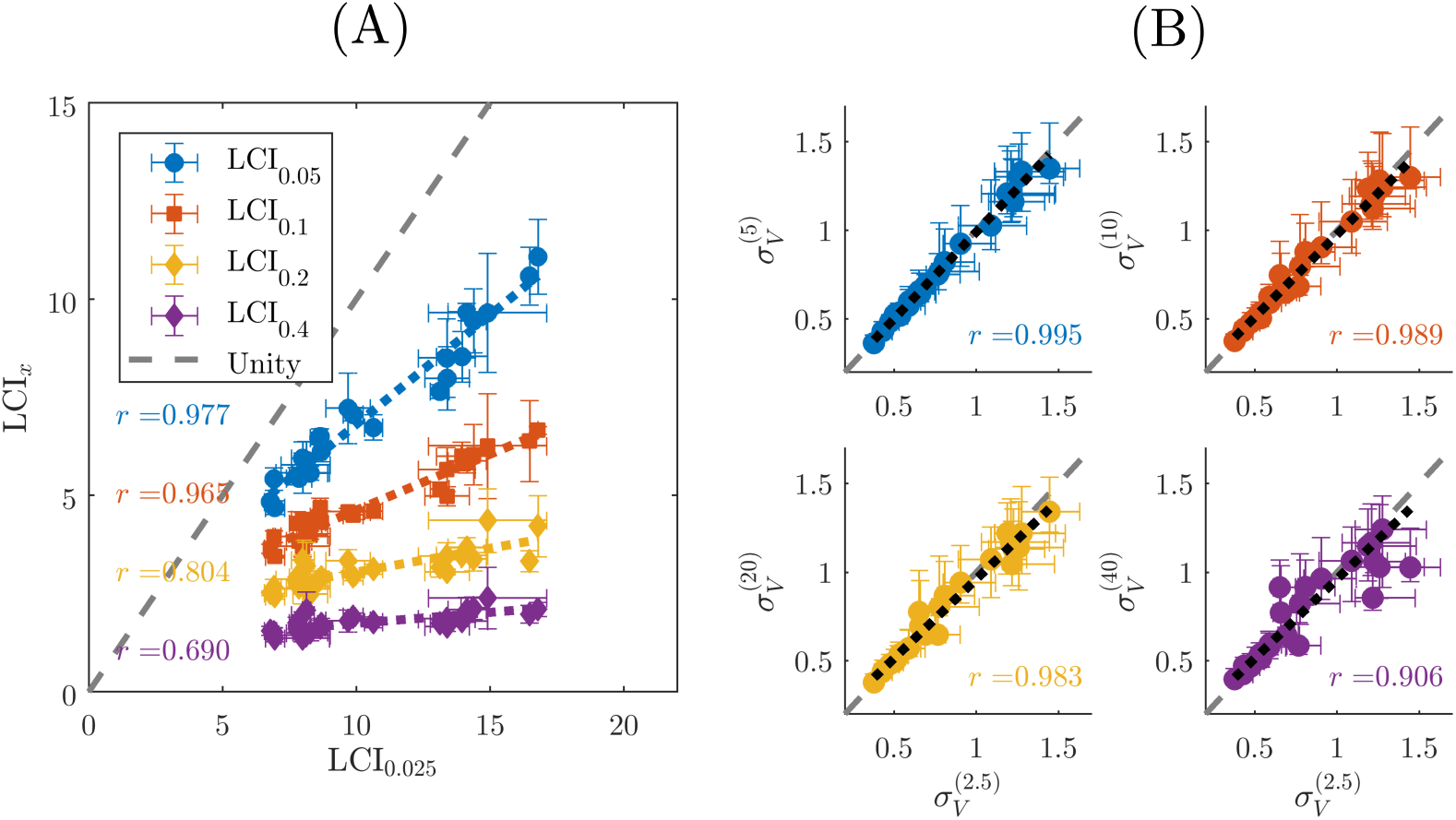
Summary of measurements from shortened MBW tests versus baseline measurements. (a) Comparison of patient LCI values (mean of 3 test repeats) at different termination thresholds (5%, 10%, 20%, and 40% of initial concentration). The error bars show the standard deviation (over test repeats) and the dotted lines show linear fits. Corresponding Pearson correlation coefficients are given next to the fit they relate to (and in the same colour). (b) Comparison of fitted (maximum a posteriori) values for the model VH parameter 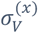 at the different termination thresholds (x=5, 10, 20, and 40% as shown by y-axes labels) plotted against 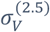 (all x-axes). Error bars show the IQR of the predicted values and the grey dashed line is the unity line. The black dotted lines show the linear fit for each plot with corresponding Pearson correlation. All p values for the linear fits shown are p < 0.001.

**Table 2:** Patient characteristics of the dataset. Age, height and weight are median values with the range in brackets, the lung function indices below are given as mean ± standard deviation. FEV_1_: forced expiratory volume in 1 second; LCI_2.5_: lung clearance index measured at the conventional endpoint of expired gas concentration 2.5% of the starting concentration; V_FRC_: lung volume at functional residual capacity; I_1/3_: fraction of pixels on MRI with low ventilation (below 1/3 of the mean); I_CV_: coefficient of variation of inhaled helium measure by image intensity.

|  | All subjects |
| --- | --- |
| Subjects (female) | 25(10) |
| Age years | 17.5 (8.9—43.7) |
| Height cm | 162 (133—189) |
| Weight kg | 55.0 (27.5—95.0) |
| FEV<sub>1</sub> % pred. | 76.5 ± 23.0 |
| FEV<sub>1</sub> z-score | -1.89 ± 1.81 |
| LCI<sub>2.5</sub> supine | 10.5 ± 3.2 |
| V<sub>FRC</sub> supine (L) | 1.50 ± 0.47 |
| I<sub>1/3</sub> | 0.146 ± 0.140 |
| I<sub>CV</sub> | 0.569 ± 0.210 |

**Table 3:** Results of truncating MBW data. The median of the washout duration per test for all subjects is given with the range in square brackets. The washout duration per test is measured for each individual by taking the mean duration of the washout period (from time of first inhalation of room air to the end of 2_nd_ exhalation following the termination threshold) over their three tests. Correlations between MRI indices of VH (I_1/3_ and I_CV_) and MBW measures (LCI and model-fitted *σ*_V_) are given in terms of their Spearman rank ρ-values, while agreement between model-predicted and measured MRI indices are given as mean bias ± 2 standard deviations.

| MBW test termination threshold | Median washout duration per test (s) | $I_{1/3}$ ρ-value | | $I_{CV}$ ρ-value | | Model agreement | |
| --- | --- | --- | --- | --- | --- | --- | --- |
| | | LCI | $\sigma_V$ | LCI | $\sigma_V$ | $I_{1/3}$ | $I_{CV}$ |
| 2.5% | 97.0 [57.3 – 215.2] | 0.82 | 0.80 | 0.75 | 0.70 | -0.003±0.118 | -0.004±0.298 |
| 5% | 73.9 [41.6 – 137.1] | 0.81 | 0.83 | 0.74 | 0.71 | 0.004±0.121 | -0.000±0.287 |
| 10% | 47.0 [32.5 – 93.6] | 0.86 | 0.82 | 0.76 | 0.72 | 0.003±0.126 | 0.005±0.291 |
| 20% | 27.9 [23.1 – 58.8] | 0.72 | 0.81 | 0.65 | 0.72 | -0.007±0.117 | -0.016±0.282 |
| 40% | 19.2 [11.0 – 38.2] | 0.59 | 0.77 | 0.51 | 0.67 | -0.017±0.154 | -0.034±0.311 |

## 4 Discussion

We have developed a method to predict the ventilation distribution in the lungs directly from MBW data. The results show good agreement with the distribution measured by hyper-polarised gas ventilation MRI (figure 4). The CF patient cohort in this study displayed varying levels of lung function, with 5/25 subjects having LCI < 8 and FEV_1_ > 90% expected, implying VH within the normal limit. Therefore, we were able to test the model across a full spectrum of LCI values. The shape of the ventilation distribution was also well characterised in the majority of cases (figure 5 and Supplementary table S1). This method therefore makes the MBW and MRI results more directly comparable. This method also has an advantage over LCI in that it incorporates the interdependence of the inferred FRC, dead-space and ventilation heterogeneity.

In addition, the model predictions of VH remained consistent as the test-data were truncated, remaining more strongly correlated with the predictions from the full dataset than LCI (figure 6 and table 3). Other studies have shown that LCI specificity and sensitivity decreases with increasing termination threshold^12^. The model we have developed is robust up to a 20% termination threshold, suggesting that washout time could be reduced by approximately 75% on average. LCI, on the other hand, proved to be consistent with MRI measures only up to a 10% termination threshold. This demonstrates that we can extract sufficient information about VH from the early breaths of washout curves to characterise the full ventilation distribution, although this requires confirmation in broader datasets. It should be noted that the prior distributions of the model parameters were still generated using estimated FRC from the full washout (*V*_*FRC*_), so the model predictions are not entirely independent of classic MBW indices, but these priors are sufficiently broad to not bias the inferred parameters. However, closed-circuit wash-in can be used to calculate more accurate priors for FRC during wash-in^34^. Thus, combining our model with closed-circuit wash-in could greatly reduce overall test time while retaining the sensitivity to VH. Even in the case of nitrogen washout, the shortened washout time would also allow a quicker recovery between tests.

This work complements the wealth of literature around predicting the ventilation distribution in the lungs from inert gas washout tests^30,31,35–40^. The underlying compartmental lung model is similar to that of previous studies including the widely used methods of Lewis et al.^36^ and the Multiple Inert Gas Washout Technique^41^. As in the more recent models^30,31^ we have used the actual MBW measured flow-rate as an input to simulations to account for the effects of variations in breath volumes (which is common in young subjects or those who have trouble regulating their breath volume). However, we designed the model to be discretised into large time-steps (10 per exhalation), improving model efficiency and enabling the use of Approximate Bayesian Computation. Bayesian inference approaches have previously only been used to extract data from the end-tidal concentrations in MBW^42^, but not to predict the ventilation distribution. Moreover, though the link between ventilation distribution and MBW curves has been studied in detail using biophysical modelling^43–46^ and benchtop experiments^47^, ours is the first study to have validated predictions of the ventilation distribution in a clinical setting using direct imaging measurement in the same patients.

The model also has some limitations. First, it assumes that gas transport to the lung units occurs in parallel, which is not a true representation of the branching airway network. Second, diffusion in the alveolar region was modelled here as instantaneous. Part of the mechanism to generate phase-III slopes^48^, particularly for gases with high molecular weight such as SF_6_, was therefore missed. This intra-acinar mixing is below the MRI resolution and furthermore the molecular weight of the gases used for the two tests are different, and so one might expect a systematic bias in VH predictions, which is not seen in figure 4. This suggests that either this cohort is dominated by convective VH or the existence of some unknown compensatory factors. Third, it is implicit in the model that the ventilation distribution is the same on inhalation and exhalation, regardless of breath volume or flow rate. These factors both affect airway closure and reopening, which may explain some of the discrepancy between predictions and outcomes. Fourth, the model in its current form is designed to model inert exogenous gases, such as SF_6_, extensions will be required to simulate exchange of N_2_ or CO_2_ as measured by certain MBW devices. Finally, an inherent difficulty of parameter identifiability occurs when VH is large enough to mean that some lung regions have very low specific ventilation, and it appears that the actual size of this unventilated region is occasionally poorly accounted for in this model (see Supplementary figure S8 for a detailed example). This leads to poorer estimates of the lung volumes (both FRC and dead-space) and greater parameter uncertainty when VH is very high, as seen in Supplementary figures S3 and S5. Related to this is the assumption of an underlying continuous distribution that is used to generate the discrete ventilation distribution in the model (in this case lognormal), which places a restrictive prior on the shapes of distribution that can be predicted (e.g. multi-modal distributions are much less likely to be predicted). Future improvements of the model will be aimed at addressing these issues and testing in larger and broader datasets.

Notwithstanding these limitations, the concept of this approach has been justified. Future refinement will be required to include more realistic acinar mixing effects and lung mechanics. To achieve this in a computationally efficient way, we may need to employ new approaches to dimensionality reduction^49^ and homogenisation^50^ of acinar and airway transport simulations. The Bayesian framework laid out here will also aid in these developments, since Bayesian model selection^32^ (which the algorithm^51^ is programmed to perform) can be used to compare the ability of different models to adequately explain the data independently of external verification.

In conclusion, our results demonstrate that this model can use an individual’s MBW test data to predict ventilation distribution in their lungs, and for the first time this been corroborated these predictions with regionally resolved ventilation imaging. This method will enable clearer interpretation of clinical data, more direct comparison between ventilation imaging and MBW data, and help to enable reductions in test time that are required to improve clinical practicality. Furthermore, by translating more abstract indices of VH (e.g. LCI or *σ*_*V*_) into more interpretable measures (e.g. the fraction of lung which is poorly ventilated *I*_1/3_) this work adds clinical value. Finally, this work is also the first example of a physiological model fitted to patient washout data using Bayesian parameter estimation, which will provide clinicians with all important estimates of uncertainty in physiological inferences.

## Data Availability

Supplementary figures and tables are linked, as well as source code used to generate the results.
Patient data (imaging and multiple-breath washout data) is not available as per the data sharing protocol of the original study upon which this work was based.

https://doi.org/10.5281/zenodo.5497647

https://github.com/CarlWhitfield/ABCModelComparison

## Acknowledgements

This report is independent research supported by the National Institute for Health Research and Health Education England and also the Medical Research Council. This work was supported by the NIHR grants; ICA-CDRF-2015-01-027 and NIHR-RP-R3-12-027 and MRC grant MR/M008894/1. CAW was supported by MRC grant MR/R024944/1. AH was supported by the NIHR Manchester Biomedical Research Centre and by NIHR grant NIHRCS12-013. The views expressed in this publication are those of the author(s) and not necessarily those of the NHS, the National Institute for Health Research, Health Education England or the Department of Health.

We would also like to acknowledge Thomas House (University of Manchester) for instrumental discussions around Bayesian methods.

## Supplementary Material

**Supplementary Text S1-S2:** Text describing methods in more detail. Text S1 describes the execution of the compartmental ventilation model. Text S2 describes the algorithm for ABC-SMC fitting.

**Supplementary Figures S1-S8:** Figures showing extra results to complement the results in the main paper. S1-S3 shows the results of parameter identifiability checks. S4 & S5 show correlations between fitted parameters and measured parameters. S6 and S7 shows examples of MBW fits and S8 shows an example MRI where MBW fitting was poor.

**Supplementary Table S1:** Measures of average agreement of the ventilation distribution predicted vs. observed in MRI, with details of how these are calculated.

## Notes

### Competing Interest Statement

The authors have declared no competing interest.

### Author Declarations

This is a predictive model validation study, no new data was collected for this study. The study upon which this work was based was approved by the Yorkshire and Humber - Leeds West Research Ethics Committee (REC reference: 16/YH/0339). Parents/guardians of children and all adult patients provided written informed consent. All data was anonymised before use in this study.

### Summary of Updates

Following multiple rounds of referee comments the structure of the article has been changed. In addition, points have been added to the introduction and discussion. However the methods and results remain substantively unchanged.

