## Supplementary Table S1 for "Model-based Bayesian inference of the ventilation distribution in patients with Cystic Fibrosis from multiple breath washout, with comparison to ventilation MRI"

### Table S1: Predicted Ventilation Distributions

For each subject, the predicted distribution (figure 4 of main text) was calculated by taking the simulated MR distribution from each accepted realisation and generating a histogram with even bin-spacing $h$ for each one. The average distribution is then constructed as the median frequency for each bin, where the medians are taken from all of the accepted realisations in the final generation of the ABC-SMC algorithm. The uncertainty in predictions is dependent on the desired precision used to characterise the distribution. We used $h=0.16$ spaced bins for the distribution measured by imaging and the distribution predicted by modelling.

Accuracy of prediction was quantified by comparing the median predicted cumulative distribution functions (CDFs) of the image intensity to those measured (note this does not depend on the bin-spacing $h$). To do this we computed the root-mean-squared residuals $\delta_{RMSE}$ of the predicted CDF against that measured for each individual. We also computed the Kolmogrov-Smirnov (K-S) statistic $\delta_{KS}$ for the two curves, defined as the maximum distance between them. In both cases this was computed by sampling each CDF at the same 1000 evenly spaced intervals. The former is a measure of the average distance between the curves (where, by their nature, the CDF functions are constrained between 0 and 1), whereas the latter is a measure of the distance between them at their point of worst agreement. To put these into context, the same measures were also computed for

D1.	A simple lognormal fit to the actual observed MRI data: We used a lognormal function with fixed unit mean (of the form in equation (S2)). To perform this, we used the maximum likelihood estimation function in matlab with an initial guess of $\sigma_{V}=0.6$.

D2.	A “null” hypothesis: The median intensity distribution predicted by the model priors (from randomly generated parameters sets).

|  | $\delta_{RMSE}$ | $\delta_{KS}$ |
| --- | --- | --- |
| Median model posterior | 0.040 (0.017) | 0.113 (0.065) |
| Mean model posterior | 0.040 (0.017) | 0.112 (0.064) |
| D1) Lognormal fit | 0.070 (0.037) | 0.156 (0.069) |
| D2) Median model prior | 0.156 (0.069) | 0.342 (0.062) |

The results are given above as the mean (s.d.) of the whole dataset.
