## Supplementary Text for "Model-based Bayesian inference of the ventilation distribution in patients with Cystic Fibrosis from multiple breath washout, with comparison to ventilation MRI"

### Supplementary Text S1: Description of algorithm for simulation a single time-step in the compartmental lung model

The volume elements in figure 3 are assumed linear in concentration and so have inert gas volume $g_{ij}= v_{ij}(c_{ij}^{\left( p \right)}+c_{ij}^{\left( d \right)})/2$. Each acinar unit $i$ exhibits instantaneous gas-mixing, so the gas concentration in the unit at time-step $n$ is $C_{i}(n)=G_{i}(n)/V_{i}(n)$, where $G_{i}$ is the volume of inert gas in the unit, and $V_{i}$ is the total gas volume of the unit. Synchronous ventilation of the units is also assumed.

At each time-step , the volume flux is perturbed by random measurement noise such that $\Delta V_{n}=\Delta V_{n}'(1+\varepsilon)$ where $\varepsilon$ is drawn from a Gaussian distribution with mean 0 and standard deviation $\sigma_{f}=0.01$ (based on upper estimates of the signal-to-noise ratio from ^66^). The algorithm for simulating each time-step is given below.

An inhalation time-step ($\Delta V_{n}>0)$ proceeds as follows (sketched in figure 3(a)):

1. A volume element with $v_{00}=\Delta V_{n}$ is added to the top of the common dead-space, with $c_{00}^{(p)}=c_{00}^{(d)}=0$.
2. Volume elements are removed from the common dead-space, starting with the distal-most element $j_{end}$up to, but not including, $j'$ such that $\sum_{j=j^{'}}^{j_{end}} v_{0j}>\Delta V_{n}$ and $V_{rem} \equiv\sum_{j=j^{'}+1}^{j_{end}} v_{0j}\leq\Delta V_{n}$.
3. The new distal-most element of the common dead-space $j'$ is split into two elements $j_{upper}$ and $j_{lower}$, such that $v_{{0j}_{lower}}=\Delta V_{n}-V_{rem}$ and $v_{{0j}_{upper}}=v_{{0j}^{'}}-v_{{0j}_{lower}}$. The interface is assigned concentration $c_{0j_{upper}}^{(d)}=c_{{0j}_{lower}}^{(p)}=(c_{0j^{'}}^{\left( p \right)}v_{j_{lower}}+c_{{0j}^{'}}^{\left( d \right)}v_{j_{upper}})/v_{j^{'}}$. Then, the element $j_{lower}$ is removed from the common dead-space.
4. All elements removed from the common dead-space in the previous two steps are replicated, the volumes scaled by $x_{i}/N_{c}$ for each lung unit $i$, and added sequentially to the proximal end of each corresponding private dead-space.
5. Elements with total volume $\Delta V_{n}x_{i}/N_{c}$ are removed from the distal end of each private dead-space $i$ using the same method as for the common dead-space in steps 2 and 3.
6. The volume $V_{i}$ of each lung unit is updated to $V_{i}\left( n \right)=V_{i}\left( n-1 \right)+ \Delta V_{n}x_{i}/N_{c}$ and the total inert gas volume in it updated to $G_{i}(n)=G_{i}(n-1)+ \sum_{j} g_{ij}$ where the sum is over the elements removed from private dead-space $i$.

An exhalation timestep ($\Delta V_{n}<0)$ proceeds as follows (sketched in figure 3(b)):

1. The volume $V_{i}$ of each lung unit is updated to $V_{i}\left( n \right)=V_{i}\left( n-1 \right)+ \Delta V_{n}x_{i}/N_{c}$ and the total inert gas volume in it is updated to $G_{i}(n)=G_{i}(n-1)+C_{i}(n-1)\Delta V_{n}x_{i}/N_{c}$.
2. A volume element $j$ with volume $v_{ij}=-\Delta Vx_{i}/N_{c}$ and concentrations $c_{ij}^{(p)}=c_{ij}^{(d)}=C_{i}(n-1)$ is added to the distal end of each private dead-space $i$.
3. Volume elements are removed from the private dead-space, starting with the proximal-most element $j=0$down to, but not including, $j'$ such that $\sum_{j=0}^{j'} v_{ij}>-\Delta V_{n}x_{i}/N_{c}$ and $V_{rem} \equiv\sum_{j=0}^{j^{'}-1} v_{ij}\leq-\Delta V_{n}x_{i}/N_{c}$.
4. The new proximal-most element $j'$ of each private dead-space $i$ is split into two elements $j_{upper}$ and $j_{lower}$, such that $v_{{ij}_{upper}}=-\Delta V_{n}x_{i}/N_{c}-V_{rem}$ and $v_{{ij}_{lower}}=v_{ij^{'}}-v_{ij_{upper}}$. The interface is assigned concentration $c_{ij_{upper}}^{(d)}=c_{ij_{lower}}^{(p)}=(c_{ij^{'}}^{\left( p \right)}v_{ij_{lower}}+c_{ij^{'}}^{\left( d \right)}v_{ij_{upper}})/v_{ij^{'}}$. Then, the element $j_{upper}$ is removed from the private dead-space $i$.
5. The elements removed from the private dead-spaces in steps 3 and 4 are combined and re-discretised as follows.
   a. There are $M\equiv\left\lfloor-\Delta V_{n}/v_{max} \right\rfloor$ new elements created with volume $v_{max}$ and a final element $j^{*}$ with volume $v_{ij^{*}}=-\Delta V_{n}-Mv_{max}$.
   b. For each new element ($j=j^{*}-M,\ldots,j^{*}$), the concentration is interpolated from the elements removed from the private dead-spaces in steps 3 and 4 such that $c_{ij}^{(d)}=c_{ij+1}^{(p)}=\sum_{i} x_{i}c_{i}^{(interp)}(jv_{max})/N_{c}$ and $c_{i}^{\left( interp \right)}\left( x \right)=\left[ \left( x-\sum_{j=0}^{j_{1}-1} v_{ij} \right)c_{ij_{1}}^{(d)}+\left( \sum_{j=0}^{j_{1}} v_{ij}-x \right)c_{ij_{1}}^{(p)} \right]/v_{ij1}$ where $j=0$ is the index of the proximal-most element ejected from dead-space $i$ and $j_{1}$ is the element such that $\sum_{j=0}^{j_{1}-1} v_{ij}\leq x$ and $\sum_{j=0}^{j_{1}} v_{ij}>x$.
6. The new elements created in step 5 are sequentially added to the distal end of the common dead-space.
7. Elements with total volume $-\Delta V_{n}$ are removed from the proximal end of the common dead-space using the same method as for the private dead-spaces in steps 3 and 4 and discarded.

After each time-step n of the simulation, concentration measurements are taken at the distal end of the common dead-space $\gamma_{n}=c_{0j^{'}}\left( n \right)+\eta$, where j’ is the distal-most element of the common dead-space, and $\eta\sim N(0,\sigma_{c})$ is measurement noise where $\sigma_{c}$ = $2\times{10}^{-4}$% SF_6_ based on Horsley et al. 2007^6^. Three model parameters are fitted which determine the ventilation distribution and resulting washout signal (V_0_, V_D_, $\sigma_{V}$).

Following MBW simulations, the same model is used to simulate hyperpolarised gas MRI measurement. A single inhalation step is simulated of size $V_{\mathrm{bag}}$ (corresponding to the volume of the bag of ${}^{3}$He and N${}_{2}$ mixture used in the MRI imaging) with unit concentration. We assume that the majority of dead-space has been successfully excluded by the airway imaging mask, and take a volume-weighted sample of the concentration in the lung units for 1000 representative datapoints, such that $I_{s}=C_{i_{s}}+\delta$ where $i_{s}$ satisfies $\sum_{i=1}^{i_{s}-1} V_{i}\leq r_{s}\sum_{i=1}^{N_{c}} V_{i}<\sum_{i=1}^{i_{s}} V_{i}$ and $r_{s}\sim U(0,1)$ for $s=1,\ldots,1000$. This means that average number of samples taken of unit $s$ is proportional to the fraction of volume this unit takes up at end-inspiration. The measurement noise $\delta\sim N(0, \sigma_{I})$ is chosen to equate to a signal-to-noise ratio of 50 such that $\sigma_{I}=0.02\sum_{i=1}^{N_{c}} G_{i}/\sum_{i=1}^{N_{c}} V_{i}$. Finally, the measurements are normalised to have unit mean (since absolute pixel intensity values are meaningless), so $\bar{I}_{s}=I_{s}/\sum_{s} I_{s}$.

### Supplementary Text S2: ABC-SMC algorithm

We define $t$ as the current `generation’ index, where a generation of simulations in one iteration of the simulation acceptance algorithm, the maximum number of generations before the algorithm terminates is $N_{t}$. Within a generation, the accepted simulations are indexed $m$, and once $N_{m}$ have been accepted, that generation of simulations is completed. We use the shorthand $\theta=\{V_{0},V_{D},\sigma_{V}\}$ to represent a parameter set, with $\theta_{t}^{(m)}$ representing the $m$th accepted parameter set in generation $t$. The model prior probability distributions (henceforth “priors”) are labelled $\pi(\theta)$, while $K_{t}({\theta|\theta}^{*})$ is the perturbation kernel, $d(\mathbf{a},\mathbf{b})$ is the distance function, and $\epsilon_{t}$ the distance threshold for generation $t.$

The algorithm proceeds as follows:

A1. Initialise $t=0$.

A2. Initialise $m=1$.

A3. Set $\epsilon_{t}$. If $t=0$, go to (a), otherwise go to (b)

(a) Draw a parameter set $\theta^{**}$ independently from the model priors $\pi(\theta)$.

(b) Draw a parameter set $\theta^{*}$ from the previous generation of accepted simulations $\{\theta_{t-1}^{\left( m \right)}\}$ with weights $\{w_{t-1}^{\left( m \right)}\}$. Next, perturb $\theta^{*}$ to get $\theta^{**}\sim K_{t}(\theta|\theta^{*})$.

A4. If $\pi(\theta^{**})=0$, return to step A3, else continue to A6.

A5. Simulate candidate dataset $\mathbf{c}^{*}=f(\theta^{**})$. If $d(\mathbf{c}^{*},\mathbf{c})>\epsilon_{t}$ return to step A3, else continue to A6.

A6. Set $\theta_{t}^{(m)}=\theta^{**}$ and calculate weight $w_{t}^{(m)}$ given by

$$\begin{matrix} w_{t}^{(m)}=\frac{\pi(\theta_{t}^{(m)})}{\sum_{m^{'}} w_{t-1}^{\left( m^{'} \right)}K_{t}(\theta_{t}^{\left( m \right)}|\theta_{t-1}^{(m')})}. \end{matrix}$$

A7. If $m<N_{m}$, set $m=m+1$ and return to step A3.

A8. If $t<N_{t}$ set $t=t+1$ and return to step A2.

The number of accepted simulations required per generation was set to $N_{m}=1120$, and the number of iterative generations $N_{t}$ was determined adaptively, such that the algorithm terminated when at least $56,000$ simulations (i.e. 1 in 50 acceptance rate) were required in the last generation. This termination threshold was implemented after finding, in a test dataset, that different MBW series required a wide range in numbers of generations to converge on a posterior distribution. The distance function $d(\mathbf{a},\mathbf{b})$ is simply taken as the root-mean-squared error between the vectors $\mathbf{a}$ and $\mathbf{b}$. We also chose the distance threshold $\epsilon_{t}$ adaptively, such that $\epsilon_{t}$ is the 60th percentile of the distances associated with the accepted parameter sets from the previous generation. Finally, we use a Gaussian kernel $K_{t}\left( {\theta|\theta}^{*} \right)\mathcal{= N(}\theta;\mu,\sigma)$where $\mu=\theta^{*}$, and $\sigma^{2}=\frac{2\sum_{m} \left( \theta_{t-1}^{\left( m \right)}-\bar{\theta}_{t-1} \right)^{2}}{N_{c}}$, where $\bar{\theta}_{t-1}=\frac{\sum_{m} \theta_{t-1}^{\left( m \right)}}{N_{c}}$.
