## Supplementary Figures for "Model-based Bayesian inference of the ventilation distribution in patients with Cystic Fibrosis from multiple breath washout, with comparison to ventilation MRI"

### Figure S1: Effect of MBW down-sampling parameters

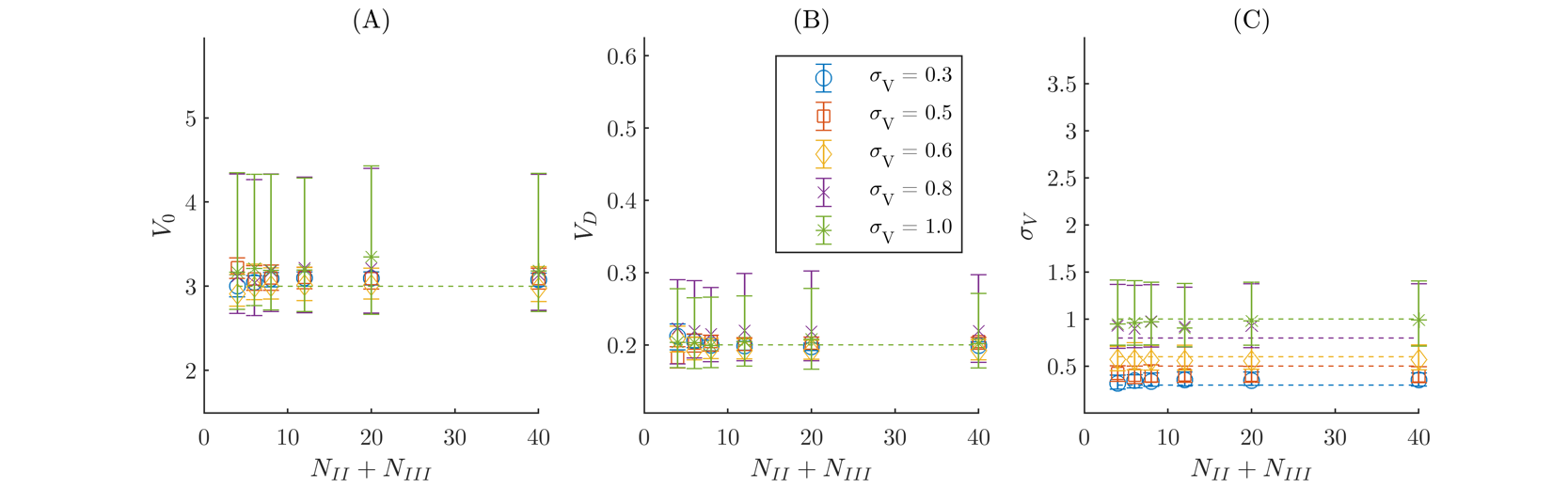

We generated artificial MBW data using realisations of the compartmental lung model with different values of $\sigma_{V}$. Data was generated using a model with $V_{0}=3$L, $V_{D}=200$ml and $\sigma_{V}=\{0.3, 0.5, 0.6, 0.8, 1.0\}$, and the tidal volume used was assumed to be a periodic step function with period 5s (2.5s inhalation and 2.5s exhalation). The tidal volumes were generated uniformly at random within the range 0.72 to 1.08L, to simulate variations in breath volume. Measurements (with simulated measurement error as described in the text) were generated for 10ms intervals and these were then outputted to match the format of the raw MBW measurements from experiments. These were then processed and fitted with the same method outlined in the main paper, but the values of $N_{II}$ and $N_{III}$ (the number of sample points used to represent phase-II and III) were varied. We set $N_{II}=N_{III}$ here to reduce the parameter space, and note that $N_{II}+N_{III}=8$ is the value used throughout the main paper. (A) – (C) shows the posterior predicted parameters for $V_{0}$, $V_{D}$, and $\sigma_{V}$ respectively as we change the number of sample points. Evidently there is very little change in the parameters recovered for $N_{II}+N_{III} \geq12$, and our choice of $N_{II}+N_{III}=8$ appears to recover the parameters well with similar uncertainty to the highest sampling-rate tested ($N_{II}+N_{III}=40$). The different markers represent fits to data generated using the $\sigma_{V}$ values listed in the legend, and the dashed lines indicate the actual parameters used to generate the data. The y-axis range is set to match the range of the model priors, and the error bars indicate the 95% prediction interval.

### Figure S2: Effect of number of model compartments

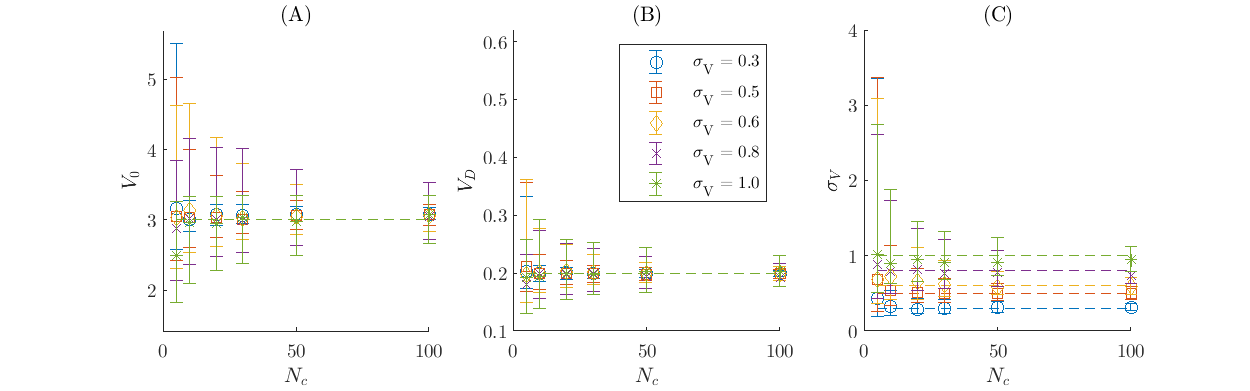

We generated artificial MBW data using realisations of the compartmental lung model with 500 compartments ($N_{c}=500)$, using the same parameters and methods as outlined in figure S2. (A) – (C) shows the posterior predicted parameters for $V_{0}$, $V_{D}$, and $\sigma_{V}$ respectively as we change the number of compartments in the fitting model. It is clear that as $N_{c}$ is increased, parameter uncertainty decreases. In the paper we use $N_{c}=50$ as a balance between execution time and reducing unnecessary uncertainty. The different markers represent fits to data generated using the $\sigma_{V}$ values listed in the legend, and the dashed lines indicate the actual parameters used to generate the data. The y-axis range is set to match the range of the model priors, and the error bars indicate the 95% prediction interval.

### Figure S3: Parameter recovery for model-generated data

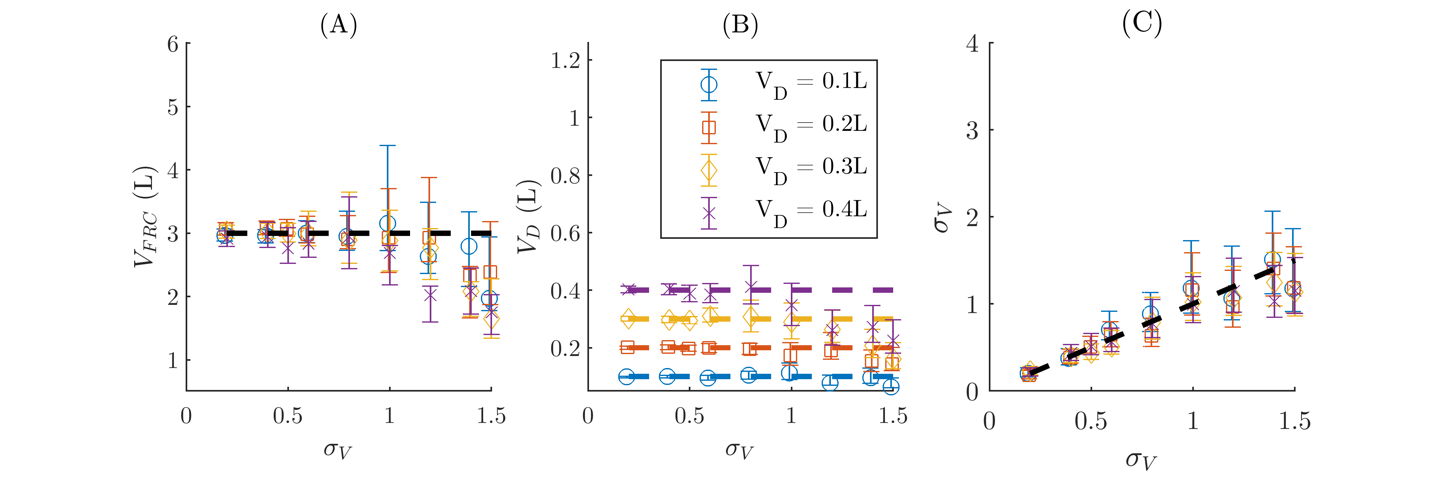

### Parameters recovered using ABC-SMC to fit simulated MBW data using realisations of the compartmental lung model with inputs $\mathbf{V}_{\mathbf{0}}\mathbf{=3}$L, $\mathbf{V}_{\mathbf{D}}\mathbf{=}\left\{ \mathbf{0.1,0.2,0.3,0.4} \right\}\mathbf{L}$ and $\boldsymbol{\sigma}_{\mathbf{V}}\boldsymbol{=\{0.3, 0.5, 0.6, 0.8, 1.0,1.5\}}$. The tidal volume used was generated uniformly at random within the range 0.72 to 1.08L for each breath, and with flow-rate in the form of a step function with period 5s (2.5s inhalation and 2.5s exhalation). Measurements (with simulated measurement error as described in the section 2.4 of the main text) were generated for 10ms intervals and these were then outputted to match the format of the raw MBW measurements from experiments. These were then processed and fitted with the same method outlined in section 2.3 in the main text. Recovered (A) $\mathbf{V}_{\mathbf{0}}$, (B) $\mathbf{V}_{\mathbf{D}}$, and (C) $\boldsymbol{\sigma}_{\mathbf{V}}$ are plotted (MAP $\boldsymbol{\pm}$ 95% PI) against input $\boldsymbol{\sigma}_{\mathbf{V}}$, with the different line colours representing the different dead-space volumes. The dashed lines indicate the input parameter values. The y-axis range was chosen to show the full extent of the prior distributions.

### Figure S4: Agreement of physiological model parameters and pulmonary function measurements

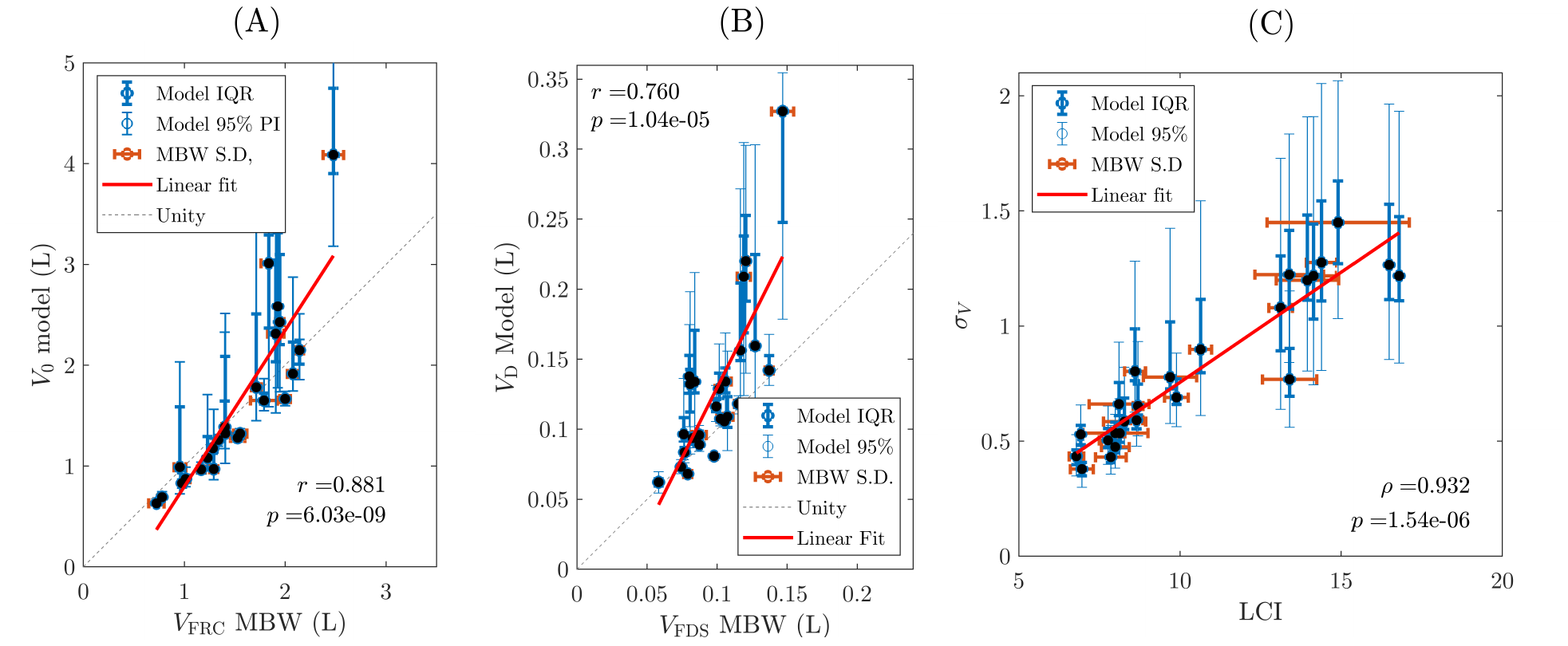

Comparison of model parameters and MBW counterparts. (A) Model *FRC versus* MBW measured FRC*.* (B) Model dead-space versus MBW measured Fowler dead-space (measured from CO_2_ trace). (C) Model VH parameter $\sigma_{V}$ vs. LCI. Key: MAP = Maximum a posteriori (most likely measurement value from ABC-SMC algorithm), IQR = Interquartile range (central 50% of sampled posterior distribution from ABC-SMC algorithm), 95% range (central 95% of sampled posterior distribution from ABC-SMC), LOA = limits of agreement (2 standard deviations either side of the mean), and MBW S.D. is the standard deviation of all MBW test repeats performed.

### Figure S5: Relationship between inferred parameters

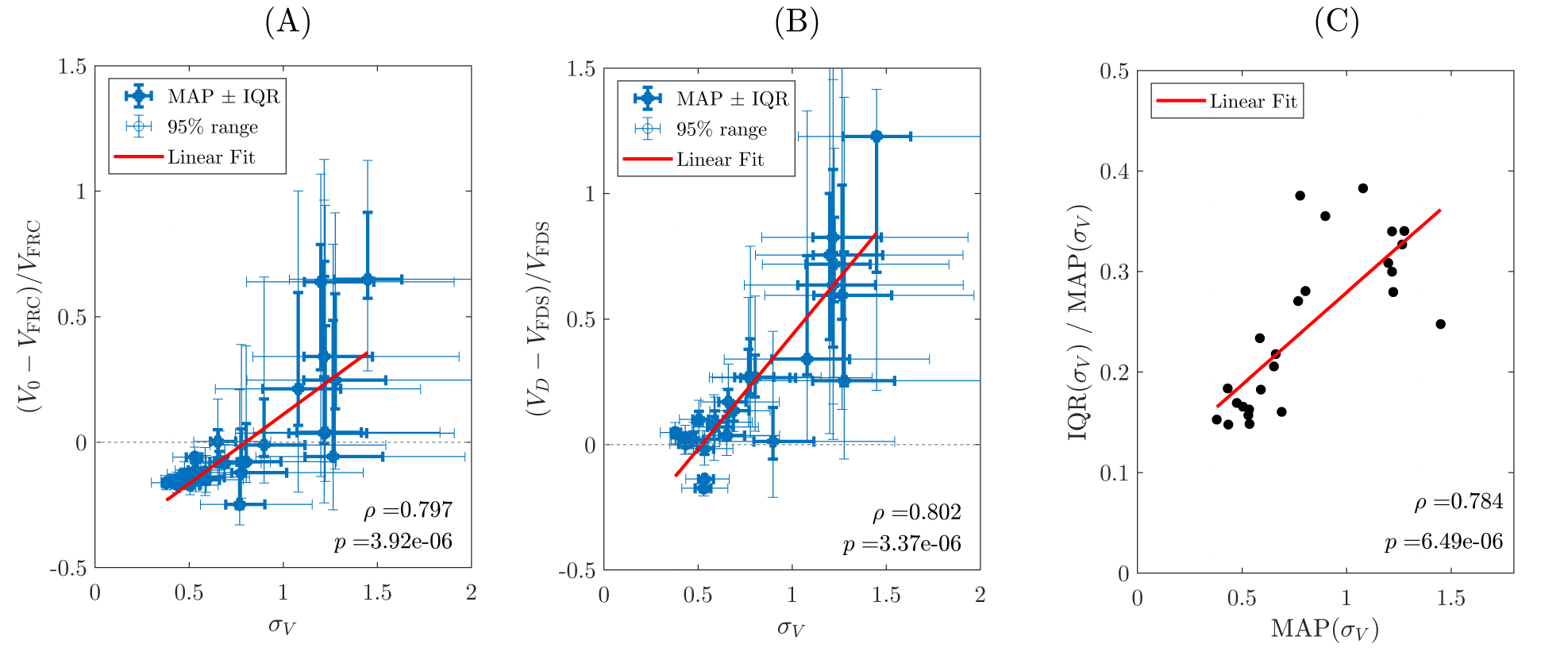

(A) The relative difference between model-predicted FRC volume and that measured by MBW vs. the ventilation heterogeneity parameter $\sigma_{V}$. (B) The relative difference between predicted dead-space volume and Fowler dead-space measured by MBW vs. the ventilation heterogeneity parameter $\sigma_{V}$. (C) The inter-quartile range (IQR) of the parameter $\sigma_{V}$ (a measure of the uncertainty) vs. the maximum a posteriori (MAP) value (i.e. the predicted value). The 95% range in plots (A) and (B) shows the limits of the central 95% of the posterior distribution for each parameter (the error in MBW measured quantities is not included in the error bars as there are insufficient experimental samples to determine the IQR or 95% range for these).

### Figure S6: Example fitted MBW traces

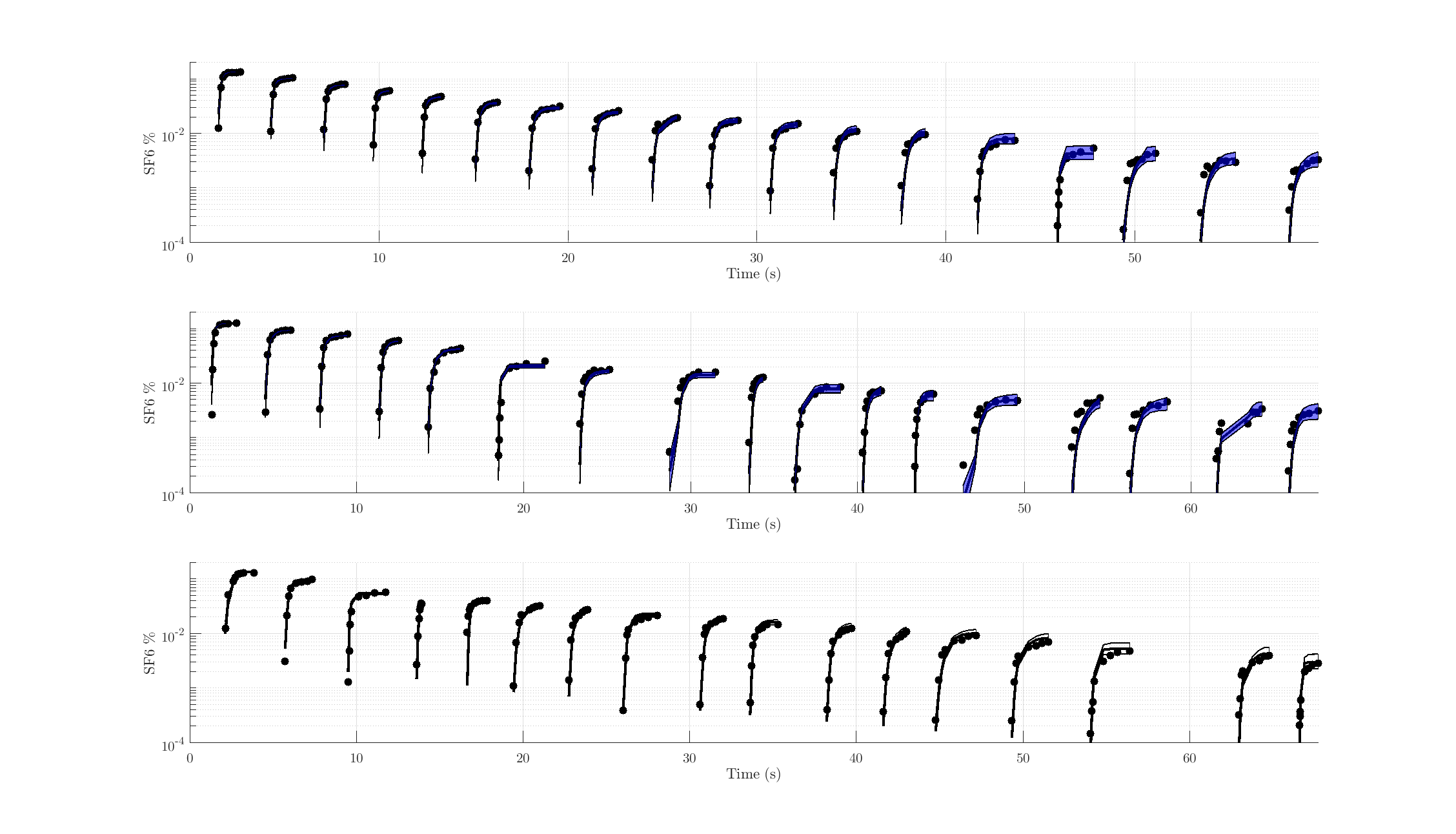

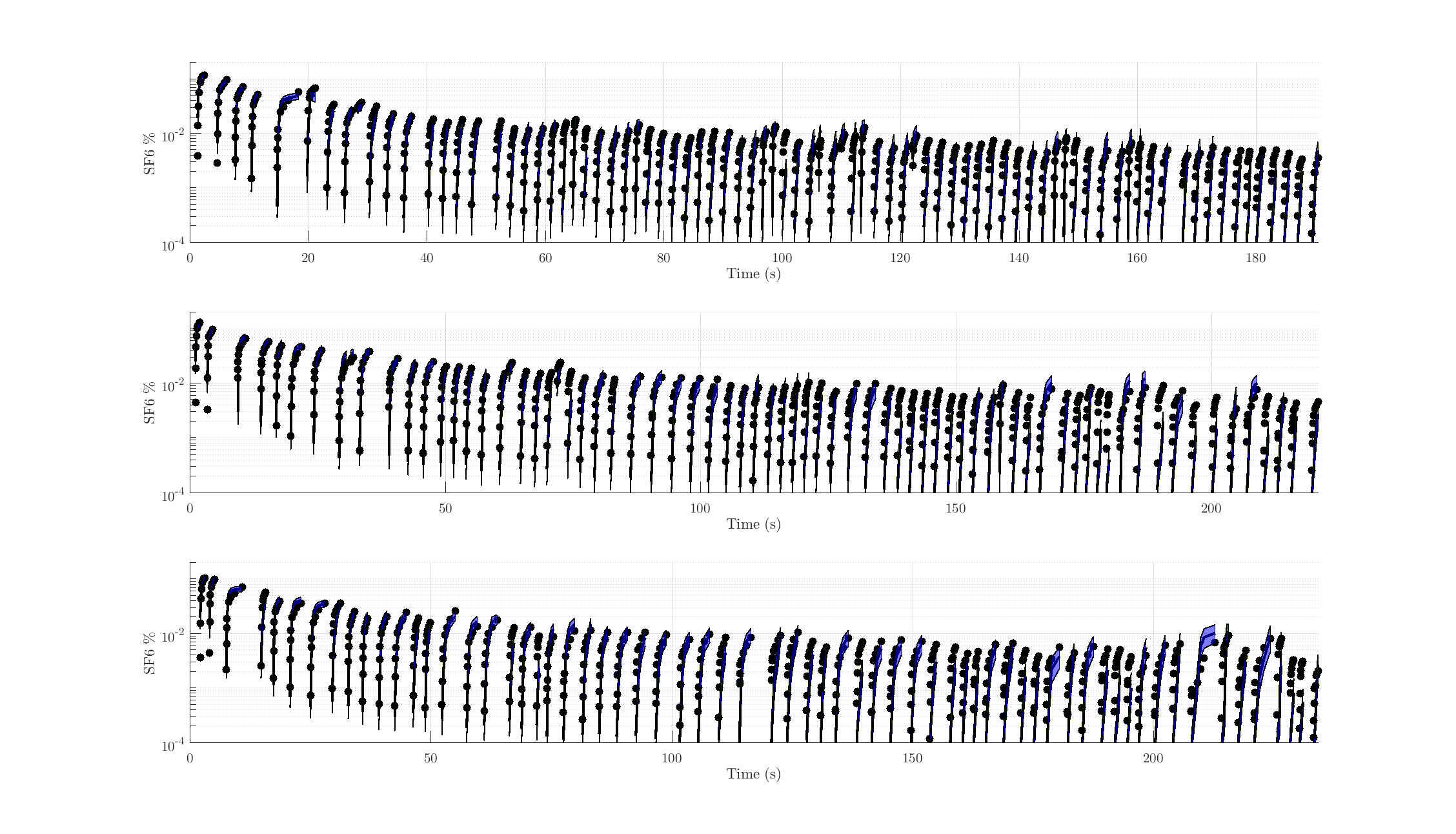

Two examples of fitted MBW traces corresponding to the patients in figures 6(b) and 6(u) respectively (i.e. the best and worst fits to the MRI data respectively). Datapoints fitted to are plotted on a log scale, the fitted line indicates the median model fit, while the shaded region is the 95% confidence interval.

### Figure S7: Example fitted MBW concentrations for different termination thresholds

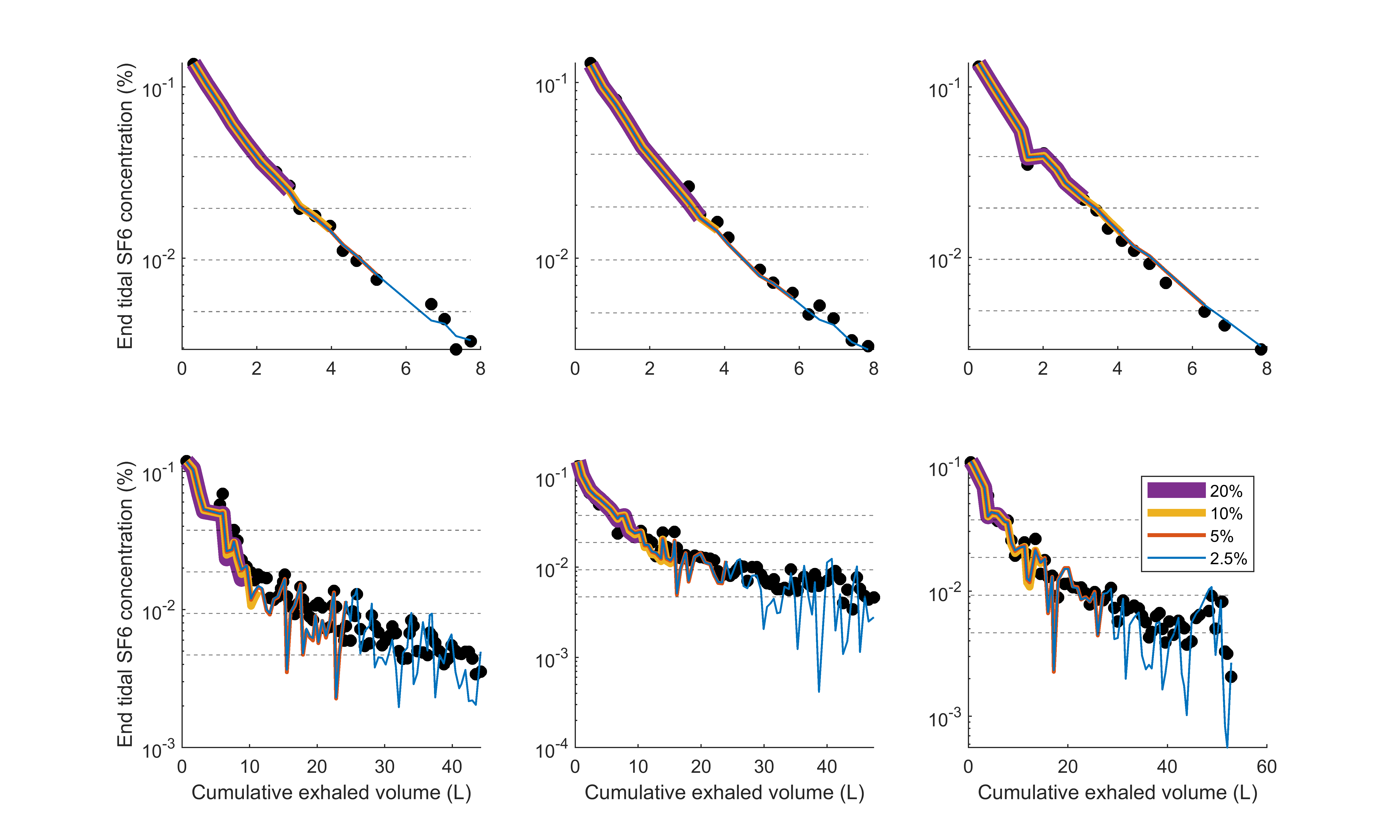

Two examples of the recorded end-tidal concentrations from repeated MBW tests (left-to-right) corresponding to the patients in Top: figure 6(b) and Bottom: figure 6(u). The median model fits to the data truncated by different MBW thresholds are shown by the solid lines as labelled. Note that the uncertainty associated with these fits is not shown for visual clarity.

### Figure S8: Example outlier case

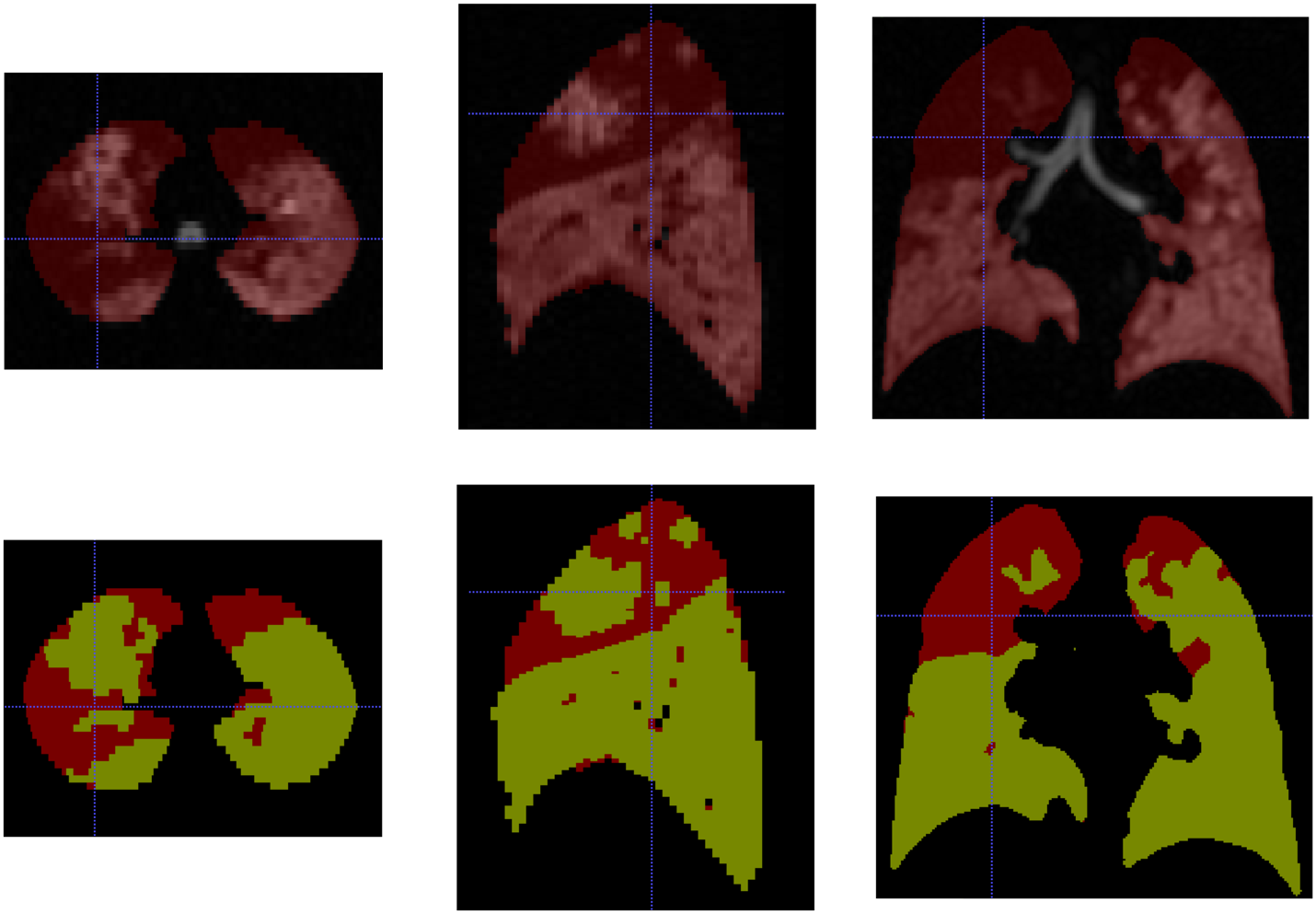

In figure 6(k) the model predicted ventilation distribution misses the spike at very low specific ventilation. The figure below shows the masked ventilation MRI image for this case (top row axial, coronal and sagittal slices from left to right), and also highlights the unventilated fraction (bottom row, same images with ventilated regions in yellow and unventilated in red). We can clearly see that the right-upper lobe (left-hand-side of coronal slice image) is barely ventilated, and that the cut-off and the lobar boundary is very clear, suggesting a proximal airway is almost completely blocked, cutting off most of this lobe from the rest of the lung, and making it hard to detect via MBW. This highlights a shortcoming of this particular model, and MBW more generally, in its inability to detect regions with extremely low specific ventilation, since, by definition, they do not contribute much to the inert gas signal during washout.
